# Serology confirms SARS-CoV-2 infection in PCR-negative children presenting with Paediatric Inflammatory Multi-System Syndrome

**DOI:** 10.1101/2020.06.05.20123117

**Authors:** Marisol Perez-Toledo, Sian E. Faustini, Sian E. Jossi, Adrian M. Shields, Hari Krishnan Kanthimathinathan, Joel D. Allen, Yasunori Watanabe, Margaret Goodall, David C. Wraith, Tonny V. Veenith, Mark T. Drayson, Deepthi Jyothish, Eslam Al-Abadi, Ashish Chikermane, Steven B. Welch, Kavitha Masilamani, Scott Hackett, Max Crispin, Barnaby R Scholefield, Adam F. Cunningham, Alex G. Richter

**Affiliations:** Institute of Immunology and Immunotherapy, University of Birmingham, Birmingham, B15 2TT, U.K.; Birmingham Clinical Trials Unit, University of Birmingham, Birmingham, B15 2TT, U.K.; Paediatric Intensive Care Unit, Birmingham Women’s and Children’s NHS Foundation Trust, Birmingham, B4 6NH, U.K.; School of Biological Sciences, University of Southampton, Southampton S017 1BJ, U.K.; Oxford Glycobiology Institute, Department of Biochemistry, University of Oxford, Oxford 0X1 3QU, U.K.; Department of Critical Care Medicine, University Hospitals Birmingham NHS Trust, Birmingham, B15 2TH, UK; Department of General Paediatrics, Birmingham Women’s and Children’s NHS Foundation Trust, Birmingham, B4 6NH, U.K.; Childhood Arthritis and Rheumatic Diseases Unit, Birmingham Women’s and Children’s NHS Foundation Trust, Birmingham, B4 6NH, U.K.; Department of Cardiology, Birmingham Women’s and Children’s NHS Foundation Trust, Birmingham, B4 6NH, U.K.; Department of Paediatrics, Birmingham Chest Clinic and Heartlands Hospital, University Hospitals Birmingham, Birmingham, B9 5SS, U.K.; West Midlands Immunodeficiency Centre, Heartlands Hospital, University Hospitals Birmingham, Birmingham, B9 5SS, U.K.; Institute of Inflammation and Ageing, University of Birmingham, Birmingham, B15 2TT, U.K.

## Abstract

**Background:** During the COVID-19 outbreak, reports have surfaced of children who present with features of a multisystem inflammatory syndrome with overlapping features of Kawasaki disease and toxic shock syndrome – Paediatric Inflammatory Multisystem Syndrome-temporally associated with SARS-CoV-2 pandemic (PIMS-TS). Initial reports find that many of the children are PCR-negative for SARS-CoV-2, so it is difficult to confirm whether this syndrome is a late complication of viral infection in an age group largely spared the worst consequences of this infection, or if this syndrome reflects enhanced surveillance.

**Methods:** Children hospitalised for symptoms consistent with PIMS-TS between 28 April and 8 May 2020, and who were PCR-negative for SARS-CoV-2, were tested for antibodies to viral spike glycoprotein using an ELISA test.

**Results:** Eight patients (age range 7–14 years, 63% male) fulfilled case-definition for PIMS-TS during the study period. Six of the eight patients required admission to intensive care. All patients exhibited significant IgG and IgA responses to viral spike glycoprotein. Further assessment showed that the IgG isotypes detected in children with PIMS-TS were of the IgGl and lgG3 subclasses, a distribution similar to that observed in samples from hospitalised adult COVID-19 patients. In contrast, lgG2 and lgG4 were not detected in children or adults. IgM was not detected in children, which contrasts with adult hospitalised adult COVID-19 patients of whom all had positive IgM responses.

**Conclusions:** Strong IgG antibody responses can be detected in PCR-negative children with PIMS-TS. The low detection rate of IgM in these patients is consistent with infection having occurred weeks previously and that the syndrome onset occurs well after the control of SARS-CoV-2 viral load. This implies that the disease is largely immune-mediated. Lastly, this indicates that serology can be an appropriate diagnostic tool in select patient groups.

## Introduction

In adults, SARS-CoV-2 virus causes respiratory infections characterised by a markedly elevated fatality rate, similar to those observed during pandemic influenza outbreaks. Those at risk of severe disease or death include the elderly, certain ethnicities and those with underlying co-morbidities such as cardiovascular disease or obesity(1). In contrast, there is a low rate of symptomatology associated with infection in children and a substantially lower risk of death(2). Nevertheless, in recent weeks reports have appeared describing rare presentations of a novel multisystem inflammatory syndrome with overlapping features of Kawasaki disease and toxic shock syndrome in children (Paediatric Inflammatory Multisystem Syndrome temporally associated with SARS-CoV-2 pandemic (PIMS-TS)), associated with SARS-CoV-2 infection(3). Diagnosis is complicated by the inconsistent detection of virus in these patients. Thus, PIMS-TS may be due to the virus or could be incidental to improved surveillance resulting from the pandemic.

Serological tests for anti-viral antibodies have not been useful to date in the immediate diagnosis of active COVID-19 infection, which relies on viral detection by PCR in conjunction with clinical presentation. This is largely due to the 7–14 day lag between infection and the development of specific antibodies. In primary infections, adaptive immunity develops with slower kinetics than on subsequent exposure. For antibody responses, IgM responses develop first, before eventually waning and IgG responses dominating thereafter. Thus, high levels of IgG in the absence of IgM are typically suggestive of infection weeks or even months previously.

Below, we present findings demonstrating that children with PIMS-TS, who are PCR-negative for SARS-CoV-2, can present with very high levels of IgG antibody to the virus.

## Materials and Methods

### Ethics statement

The patients’ samples were either tested as part of routine diagnostics on in house COVID-19 antibody ELISAs run by the UKAS accredited Clinical Immunology Service at the University of Birmingham or used for assay development. The ethical approval for this work and the use of these samples was provided by the awarding bodies of the University of Birmingham Research Ethics Committee, the South Birmingham Research Ethics Committee and the National Research Ethics Service Committee West Midlands. All approvals are overseen by the United Kingdom National Health Service and this is therefore a NHS Health Research Authority approved study. All patients and/or their parents/legal guardians provided signed informed consent to inclusion of de-identified data in this report.

### Patient cohort and samples

We used a case definition consistent with Royal College of Paediatrics and Child Health guidelines and patients were identified based on fulfilling the case definition for PIMS-TS described in Table 1. All were admitted to hospital between 28^th^ April-8^th^ May 2020. Tests for SARS-CoV-2 infection by PCR gave negative results. All patients received standard supportive care that included empirical antibiotics, respiratory and cardiovascular support as indicated. Patients received intravenous immunoglobulin and/or steroids if they fulfilled either full or atypical Kawasaki disease criteria.

**Table 1.**
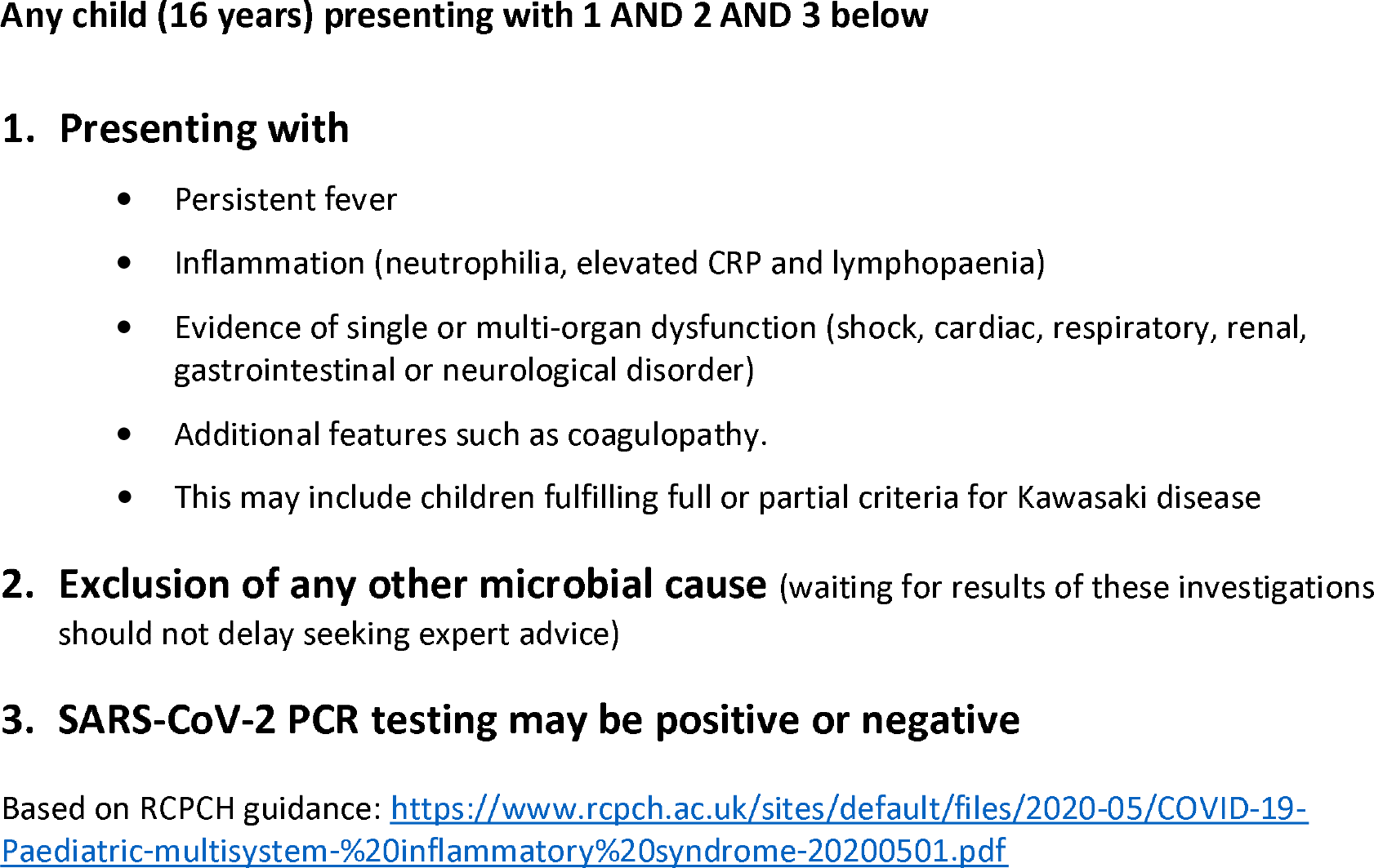
Case definition for Paediatric Inflammatory Multisystem Syndrome temporally associated with SARS-CoV-2 pandemic.

### ELISA to SARS-CoV-2 spike glycoprotein

Antibodies to near-full-length trimeric viral spike glycoprotein(4, 5), were detected by ELISA. High-binding plates (Greiner Bio-One) were coated with spike glycoprotein (1 μg/ml) and blocked with with Stabilcoat solution (Sigma Aldrich) before test serum was added at 1:40 or 1:50 and diluted 5-fold down the plate. HRP-labelled mouse monoclonal anti-human IgG, IgA IgM, IgG_1-4_ secondary antibodies, generated at the University of Birmingham (available from Abingdon Health Ltd), were added individually or combined and HRP activity detected using TMB core (Bio-rad).

## Results

### Patient presentation

Eight patients, 5 (63%) male and median age 9 (range 7–14) years, fulfilled case definition for PIMS-TS and had SARS-CoV-2 serology tested. Patients were of mixed ethnicity. What was unusual was the clustering of cases and in light of recent reports, the potential that this was a SARS-CoV-2 presentation was considered. 7 patients had overlapping features of hyper-inflammation with either typical or atypical Kawasaki disease, and one patient had overlapping features of hyper-inflammation and toxic shock syndrome. All patients had fever and at least one gastrointestinal symptom (abdominal pain, vomiting and diarrhoea) and 75% had a rash. Hyperinflammation was supported by presence of fever and the median (IQR) CRP was 188 (136–255) mg/L and ferritin was 1325 (819–2121) μg/L in this cohort of children. 63% of patients had impaired myocardial function on echocardiography. 75% required admission to paediatric intensive care predominantly for cardiovascular support due to hypotension. All patients improved with supportive therapy that included immunomodulation with immunoglobulins and/or steroids and were discharged from PICU, remaining hospital inpatients.

### Antibody detection to viral spike glycoprotein

Sera from these children were tested against viral spike glycoprotein, the major immunodominant antigen and compared to pre-2019 sera and plasma from adults with severe COVID-19 infections admitted to the intensive treatment unit (ITU). A screening test to detect IgG, IgA and IgM was performed at a single dilution of 1:40 of sample, which demonstrated that all children had antibody against the SARS-CoV-2 spike glycoprotein (Fig. 1A). Since antibody isotypes can reflect recent infection (IgM), or more historic infections (IgG and IgA), we examined individual antibody isotypes. In children, IgM levels were similar to pre-2019 sera; in contrast, spike glycoprotein-specific IgM levels were high in adult ITU COVID-19 patients. (Fig. 1B). IgA and IgG were more similar in children and adult COVID-19 patients. Assessment of IgG isotypes, which informs on the effector function of the antibodies, revealed IgGl and lgG3 were the predominant isotypes present in these children and in adults (Fig. 1C), with lgG2 and lgG4 similar to negative controls in all but one child, who had a weak IgG4 response (data not shown). Therefore, children with Kawasaki-like inflammatory syndrome who are negative by PCR can have high IgGl, lgG3 and IgA antibody levels to SARS-CoV-2 in the absence of maintained IgM levels.

**Figure 1.**
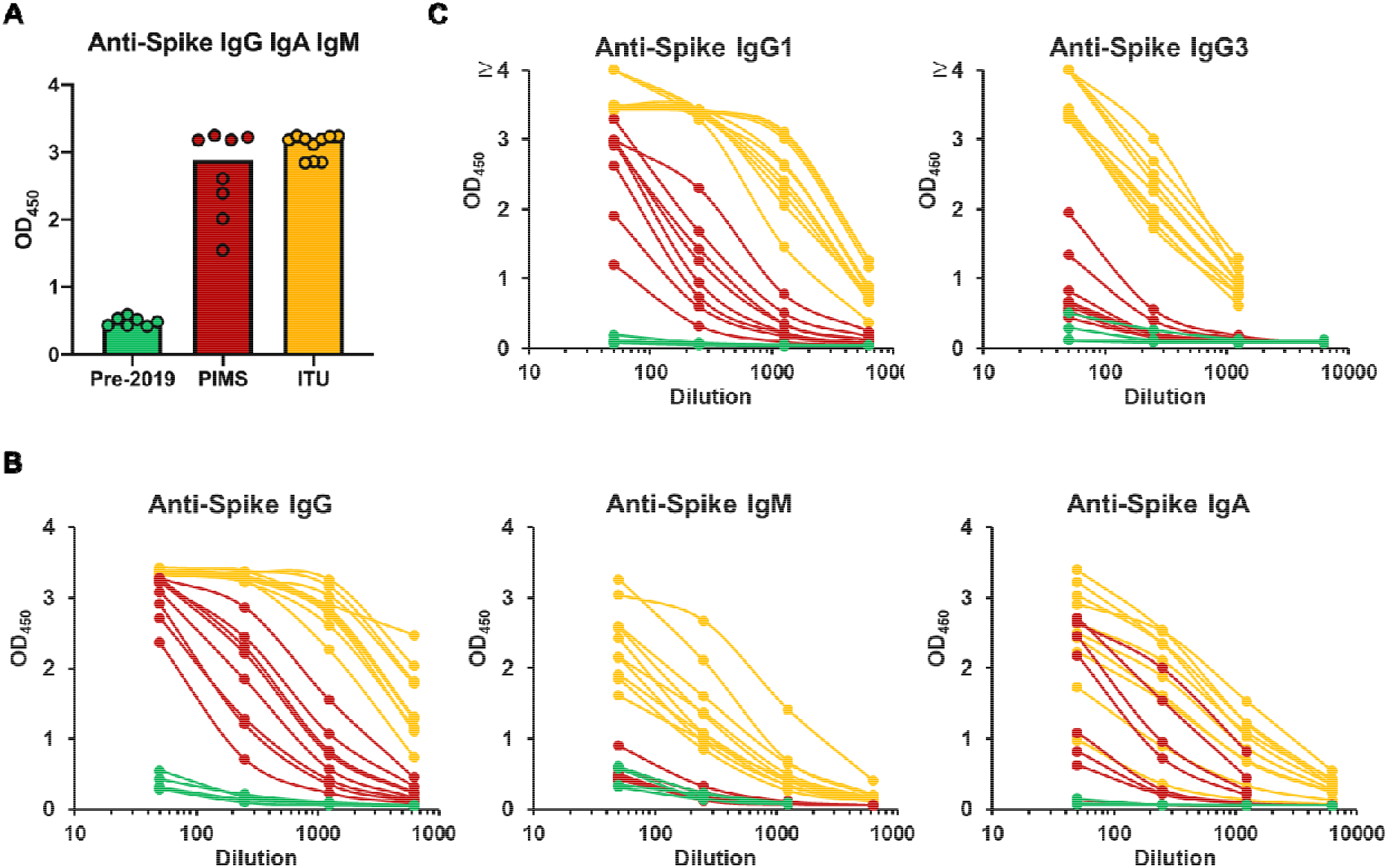
Detection of anti-SARS-CoV-2 antibody responses in children with PIMS-TS. Serological responses were detected against purified near-full-length trimeric SARS-CoV-2 viral spike glycoprotein by ELISA. **A)** Absorbance values of individual sera at a single dilution (1:40) from pre-2019 healthy adult donors (green) or sera from children with PIMS-TS (red), or plasma from adult ITU patients (orange) detected using combined HRP-labelled anti-IgG, IgA and IgM. One symbol represents results for a single serum and the bar shows the median values for each group. **B)** Absorbance values for individual sera from pre-2019 negative control donors (green), children with PIMS-TS (red), or plasma from adult ITU patients (orange) serially diluted five-fold from 1:50, primary antibodies were detected using HRP-labelled anti-IgG, IgA or IgM individually, or **C)** HRP-labelled IgGlor lgG3.

## Discussion

Recent anecdotal reports of a Kawasaki-like inflammatory syndrome, often without detection of SARS-CoV-2 virus, appear at odds with the relatively mild or asymptomatic presentation of SARS-CoV-2 infection in the vast majority of children(2, 6). Testing for SARS-CoV-2 infection in eight hospitalised children presenting with PIMS-TS found significant levels of IgG and IgA to this virus despite no evidence of current infection by PCR. Although PCR detection of infection is an imperfect technique, it is the nearest to a gold standard for determining active infection(7). Not detecting virus in any of these eight children is unlikely to be due to false-negative PCR results and more consistent with presentation after viral clearance. Compatible with this conclusion, all samples had low levels of specific IgM and high levels of IgG, indicating that infection may have occurred weeks or even months previously. As we report in a forthcoming manuscript, after testing hundreds of pre-pandemic control sera we have found minimal cross-reactivity of pre-pandemic sera with the spike glycoprotein, indicating our assay is highly specific for SARS-CoV-2 and that this virus is indeed the aetiological trigger.

The absence of current infection would suggest that symptomatology in these children relates to immune-mediated pathology. Immune-mediated disease has been suggested to contribute to disease severity in adult infections(8). Associated with this is the detection of IgGl and lgG3 in these children. These isotypes are associated with complement activation(9), which has been shown to be enhanced in adult patients(10), otherwise this could simply be a proxy for other immune activity. An alternative mechanism would be antibody-dependent enhancement (ADE) which has been reported with other coronaviruses(11–13). This is a paradoxical phenomenon in which binding of non-neutralizing antibodies to a virus enhances entry into host cells, resulting in more severe disease in secondary infections, but this is unlikely in these PCR negative children. Other routes to immune-mediated damage may also be important in these individual cases. The SARS-CoV-2 specific antibodies could either induce a pathogenic response to a self-antigen through molecular mimicry or could simply be markers of a ‘hit and run’ virus induced inflammatory condition. This report does not explain the mechanism behind the antibody pattern but it is an important observation when considering potential complications post-vaccination and warrants further investigation.

In summary, PCR-negative patients can present with a severe inflammatory syndrome whose aetiology can only be determined through antibody testing. This is important, as until now, serology has not been useful diagnostically, only for epidemiology. This therefore offers a widening of the value of serology in the identification and understanding of infections caused by SARS-CoV-2. Indeed, since all patients were positive serologically, it may be worth considering amending the definition of PIMS-TS so that TS is not just “temporally associated with SARS-CoV-2 pandemic”, but “triggered by SARS-CoV-2 infection”.

## Data Availability

The data that support the findings of this study are available from the corresponding author upon reasonable request.

## Acknowledgements

We would like to thank the University of Birmingham Clinical Immunology Service for their invaluable support in sample collection and processing. AFC is grateful for funding from The Medical Research Council and The Institute for Global Innovation, The University of Birmingham for funding. This study was supported by the UK National Institute for Health Research, Birmingham Biomedical Research Centres Funding scheme. Dr Barnaby Scholefield is funded by the NIHR Clinician Scientist fellowship programme. The work in Prof. Max Crispin’s laboratory was funded by the International AIDS Vaccine Initiative, Bill and Melinda Gates Foundation through the Collaboration for AIDS Vaccine Discovery (OPP1084519 and OPP1115782), the Scripps Consortium for HIV Vaccine Development (CHAVD) (AI144462), and the University of Southampton Coronavirus Response Fund which has over 1000 donors from around the world. We would like to acknowledge the support of the Birmingham Women’s and Children’s Hospital NHS Foundation trust staff and patients, including Drs Fiona Reynolds, Jim Gray, Mitul Patel, Phillip Hurley, Tristan Ramcharan, Habib Ali, Sakeena Samar, Penny Davis, Kathryn Harrison, William Coles, Pam Dawson, Sean Monaghan, Deevena Chinthala, Heather Duncan, Nick Richens and Sanket Sontakke. We thank Jason McLellan for the expression plasmid for the SARS-CoV-2 glycoprotein. We are grateful to Dr Galit Alter, Harvard University for helpful comments. We thank The Binding Site for technical assistance.

## References

1. Jordan RE, Adab P, Cheng KK. Covid-19: risk factors for severe disease and death. BMJ (Clinical research ed). 2020;368:mll98.

2. Ludvigsson JF. Systematic review of COVID-19 in children shows milder cases and a better prognosis than adults. Acta paediatrica (Oslo, Norway: 1992). 2020;109(6):1088–95.

3. Riphagen S, Gomez X, Gonzalez-Martinez C, Wilkinson N, Theocharis P. Hyperinflammatory shock in children during COVID-19 pandemic. Lancet 2020.

4. Watanabe Y, Allen JD, Wrapp D, McLellan JS, Crispin M. Site-specific glycan analysis of the SARS-CoV-2 spike. Science (New York, NY). 2020.

5. Wrapp D, Wang N, Corbett KS, Goldsmith JA, Hsieh CL, Abiona O, et al. Cryo-EM structure of the 2019-nCoV spike in the prefusion conformation. Science (New York, NY). 2020;367(6483):1260–3.

6. Wu Z, McGoogan JM. Characteristics of and Important Lessons From the Coronavirus Disease 2019 (COVID-19) Outbreak in China: Summary of a Report of 72H314 Cases From the Chinese Center for Disease Control and Prevention. Jama. 2020.

7. Esbin MN, Whitney ON, Chong S, Maurer A, Darzacq X, Tjian R. Overcoming the bottleneck to widespread testing: A rapid review of nucleic acid testing approaches for COVID-19 detection. RNA (New York, NY). 2020.

8. Kadkhoda K. COVID-19: an Immunopathological View. mSphere. 2020;5(2).

9. Valenzuela NM, Schaub S. The Biology of IgG Subclasses and Their Clinical Relevance to Transplantation. Transplantation. 2018;102(1S Suppl 1):S7–s13.

10. Risitano AM, Mastellos DC, Huber-Lang M, Yancopoulou D, Garlanda C, Ciceri F, et al. Complement as a target in COVID-19? Nature reviews Immunology. 2020:1–2.

11. Jaume M, Yip MS, Cheung CY, Leung HL, Li PH, Kien F, et al. Anti-severe acute respiratory syndrome Coronavirus spike antibodies trigger infection of human immune cells via a pH-and cysteine protease-independent FcγR pathway. Journal of virology. 2011;85(20):10582–97.

12. Kam YW, Kien F, Roberts A, Cheung YC, Lamirande EW, Vogel L, et al. Antibodies against trimeric S glycoprotein protect hamsters against SARS-CoV challenge despite their capacity to mediate FcgammaRII-dependent entry into B cells in vitro. Vaccine. 2007;25(4):729–40.

13. Wang SF, Tseng SP, Yen CH, Yang JY, Tsao CH, Shen CW, et al. Antibody-dependent SARS Coronavirus infection is mediated by antibodies against spike proteins. Biochemical and biophysical research communications. 2014;451(2):208–14.

